# A multi-analyte serum biomarker panel for early detection of pancreatic adenocarcinoma

**DOI:** 10.1101/2022.03.03.22271867

**Authors:** Matthew A. Firpo, Kenneth M. Boucher, Josh Bleicher, Gayatri D. Khanderao, Alessandra Rosati, Katherine E. Poruk, Sama Kamal, Liberato Marzullo, Margot De Marco, Antonia Falco, Armando Genovese, Jessica M. Adler, Vincenzo De Laurenzi, Douglas G. Adler, Kajsa E. Affolter, Ignacio Garrido-Laguna, Courtney L. Scaife, M. Caterina Turco, Sean J. Mulvihill

## Abstract

**Purpose:** We determined whether a large, multi-analyte panel of circulating biomarkers can improve detection of early-stage pancreatic ductal adenocarcinoma (PDAC).

**Experimental Design:** We defined a biologically relevant subspace of blood analytes based on previous identification in premalignant lesions or early-stage PDAC and evaluated each in pilot studies. The 31 analytes that met minimum diagnostic accuracy were measured in serum of 837 subjects (461 healthy, 194 benign pancreatic disease, 182 early stage PDAC). We used machine learning to develop classification algorithms using the relationship between subjects based on their changes across the predictors. Model performance was subsequently evaluated in an independent validation data set from 186 additional subjects.

**Results:** A classification model was trained on 669 subjects (358 healthy, 159 benign, 152 early-stage PDAC). Model evaluation on a hold-out test set of 168 subjects (103 healthy, 35 benign, 30 early-stage PDAC) yielded an area under the receiver operating characteristic (ROC) curve (AUC) of 0.920 for classification of PDAC from non-PDAC (benign and healthy controls) and an AUC of 0.944 for PDAC vs. healthy controls. The algorithm was then validated in 146 subsequent cases presenting with pancreatic disease (73 benign pancreatic disease, 73 early and late stage PDAC) as well as 40 healthy control subjects. The validation set yielded an AUC of 0.919 for classification of PDAC from non-PDAC and an AUC of 0.925 for PDAC vs. healthy controls.

**Conclusions:** Individually weak serum biomarkers can be combined into a strong classification algorithm to develop a blood test to identify patients who may benefit from further testing.

## Introduction

Pancreatic adenocarcinoma (PDAC) is one of the most highly lethal diseases resulting in over 47,000 deaths in the United States (US) annually and nearly 460,000 deaths worldwide (1,2). This number is expected to rise, as PDAC incidence rates have been increasing by 1% per year since the 1970s (3). This has led to projections that PDAC will be the second leading cause of cancer deaths by 2030 (4). Surgical resection remains the mainstay of curative therapy, with 5-year survival rising to 25% for patients who present with resectable disease (5). Advances in pancreatic surgery and the expanded use of neoadjuvant chemotherapy have increased the fraction of patients eligible for surgery and decreased operative morbidity (6-8); however, the majority of patients still present with advanced, unresectable disease for which treatments remain ineffective (9). Five year survival for PDAC remains around 10%, improved from 5% survival rates 20 years ago (10).

An accurate, cost-effective screening protocol could substantially increase the number of patients eligible for surgical resection of newly diagnosed PDAC and greatly improve current PDAC-associated oncologic outcomes. Available evidence suggests at least a 5-year period between the development of malignant founder cells and acquisition of metastatic capacity, offering a window of opportunity for identification of early-stage, potentially curable disease (11). Screening for PDAC is a controversial topic however, with no currently effective screening tool for detecting early tumors or asymptomatic tumors in the general population (12). The US Preventative Services Task Force (USPSTF) recently discouraged screening for pancreatic cancer in asymptomatic adults, as the risks of harm currently outweigh the benefits (13).

A blood assay screening for biomarkers with high sensitivity and specificity would be the ideal screening test for PDAC (5). The costs of screening with imaging are prohibitively high for the general population, with a needed dysplasia prevalence of at least 16% in the screening cohort to be considered cost-effective (14,15). A blood-based test may be useful as a primary screening tool for assessing the need for secondary screening by imaging. Limiting screening to groups with high-risk for developing PDAC would increase the pretest probability and reasonably offset the risk of false-positive diagnoses and costs with the survival benefits of early detection. Individual biomarkers, such as CEA and CA 19-9, lack the accuracy to be used independently for early-stage PDAC screening; however, a blood-based diagnostic panel relying on multiple biomarkers could be developed to improve sensitivity and specificity of a screening assay (16,17).

This study describes the performance of a multianalyte, blood-based primary screening test developed for detection of early stage PDAC in order to suitably inform the need for secondary screening using imaging. Potential analytes were assembled from a large body of prior studies that investigated circulating biomarkers in early-stage disease (18-73). We used statistical learning to develop classification algorithms using the relationship between classes based on the changes across the 31 analytes. A diagnostic algorithm was devised in a large development cohort and subsequently validated in an independent sample set.

## Materials and Methods

### Subjects and Sample Collection

Subjects presenting with suspicion of pancreatic disease as well as accompanying adults to serve as healthy control subjects were approached for enrollment in an institute-wide research protocol at the Huntsman Cancer Institute at the University of Utah. Blood was collected from consenting adults and processed for serum, aliquoted, and frozen within two hours of collection. Available samples were periodically queried for inclusion in this study and samples from all disease cases meeting the inclusion criterion were assayed. Cases were chosen for inclusion if they had cytologically confirmed stage IA, IB, IIA, or IIB PDAC (N = 182) with an available treatment-naïve serum sample. Control samples from patients with benign pancreatic disease included subjects with chronic pancreatitis (ChPT, N = 100) and intraductal papillary mucinous neoplasm (IPMN, N = 94). Samples for IPMN cases were included if the patient had three years of subsequent follow-up without intervention for progression. Healthy control samples (CON, total N = 461) were included from accompanying adults (independent of the diagnosis of the primary case) and cases obtained from the University of Salerno (N = 39). Additional control cases included excess sera obtained from a reference laboratory (ARUP Laboratories) managed by the University of Utah Department of Pathology (N = 287). Healthy control cases were sex-matched and age approximated to the PDAC cases. Subject characteristics are shown in Table 1. Informed written consent was obtained from each subject enrolled in the research protocol and all studies were performed with the approval of the Institutional Review Board at the University of Utah in accordance to the principles of the Helsinki Declaration and the U.S. Common Rule.

**Table 1:** Subject Characteristics

|  | PDAC | Controls | IPMN | Chronic Pancreatitis |
| --- | --- | --- | --- | --- |
| **<u>Development Data Set</u>** |  |  |  |  |
| Cases | 182 | 461 | 94 | 100 |
| Age, median (IQR) | 67 (61 – 75) | 57 (53 – 65) | 66 (57 – 72) | 58 (51 – 64) |
| Male Sex (%) | 105 (58) | 193 (42) | 39 (41) | 57 (57) |
| Stage |  |  |  |  |
| I (%) | 26 (14) |  |  |  |
| IIA (%) | 63 (35) |  |  |  |
| IIB (%) | 93 (51) |  |  |  |
| **<u>80% Training Set</u>** |  |  |  |  |
| Cases | 152 | 358 | 77 | 82 |
| Age, median (IQR) | 67 (60 – 75) | 57 (52 – 65) | 65 (54 – 70) | 58 (50 – 65) |
| Male Sex (%) | 87 (57) | 152 (42) | 32 (42) | 44 (54) |
| Stage |  |  |  |  |
| I (%) | 22 (14) |  |  |  |
| IIA (%) | 55 (36) |  |  |  |
| IIB (%) | 75 (49) |  |  |  |
| **<u>20% Test Set</u>** |  |  |  |  |
| Cases | 30 | 103 | 17 | 18 |
| Age, median (IQR) | 69 (64 – 73) | 58 (53 – 64) | 73 (67 – 76) | 60 (53 – 64) |
| Male Sex (%) | 18 (60) | 41 (40) | 7 (41) | 13 (72) |
| Stage |  |  |  |  |
| I (%) | 4 (13) |  |  |  |
| IIA (%) | 8 (27) |  |  |  |
| IIB (%) | 18 (60) |  |  |  |
| **<u>Validation Set</u>** |  |  |  |  |
| Cases | 73 | 40 | 41 | 32 |
| Age, median (IQR) | 69 (61 – 74) | 57 (51 – 67) | 67 (57 – 73) | 50 (39 – 62) |
| Male Sex (%) | 34 (47) | 20 (50) | 16 (39) | 17 (53) |
| Stage |  |  |  |  |
| I (%) | 17 (23) |  |  |  |
| IIA (%) | 7 (10) |  |  |  |
| IIB (%) | 8 (11) |  |  |  |
| III (%) | 19 (26) |  |  |  |
| IV (%) | 17 (23) |  |  |  |
| unknown | 5 (7) |  |  |  |

### ELISA

A total of 837 serum samples were assayed for 31 analytes by ELISA. Samples were processed in four batches over the course of 2 years. Commercially available kits (Supplementary Table S1) were used for analyte quantification according to the manufacturers recommended protocols, including primary antibody optimization, where appropriate. For analyte readings that were above the linear range of detection, samples were diluted and re-assayed. For analyte readings that were below the lowest detectable concentration, samples were re-assayed using higher volumes, if possible, or assigned a value of half the lowest detectable concentration. The assay for one analyte, BAG3, was developed in house and previously described (28,74). For three analytes (HP, LRG1, PRG4), changes in availability of commercial kits required use of different vendors. In order to adjust for changes in antibody concentration and dilution factor, correction factors were calculated based on a comparison of standards from the previous kit to standards of the new kit. These correction factors were validated on readings from control serum spiked with known concentrations of all 31 analytes designed to give readings within the linear range of detection and evaluated on both kits. The raw data was log transformed and adjusted for sex, age, and batch (fitting a natural spline with 2 degrees of freedom) using a linear model. The resulting adjusted data was normalized by centering and dividing by the standard deviation of the healthy control data.

### Data Analysis

Statistical analyses and modeling were performed using R version 3.6.1 (75). For diagnostic performance of individual analytes, areas under the receiver operating characteristic (ROC) curve (AUC) were determined. Bootstrap resampling was utilized to estimate results from repeated analyses. A random sampling with replacement of calibration samples was performed 2000 times and the range of values were then recorded and compared to the average result. Ensemble models were built using the caretEnsemble package in R (76) by stacking using gradient boosting. A non-linear method is appropriate given the large data set, individual models yielding similar accuracies, and different models acting on different subsets of the data. For modeling, all data with known class in the 837-sample data set were adjusted together before randomly separating into training (80%) and test (20%) sets. Discriminate algorithms were developed from the training data set for dichotomous classification of controls (including healthy, chronic pancreatitis, and benign IPMN subjects) and early stage PDAC cases. Using the training data set, models were built to optimize area under the ROC curve, emphasizing overall model accuracy. Method selection and tuning parameters for individual modeling methods were evaluated in an intermediate data subset consisting of 74 healthy control, 60 chronic pancreatitis, 76 IPMN, and 122 early-stage PDAC cases. Individual model performance was assessed by 10-fold cross validation. Ensemble classification models involved first optimizing submodels using individual classification methods (‘glmnet’, ‘svmRadial’, ‘rf’, ‘nnet’, ‘knn’ packages in R) and then optimizing classification by combining the individual methods by stacking (gbm package in R). Again, performance of the ensemble models was assessed by 10-fold cross validation. Ensemble classification models were then used to predict class probabilities to the samples in the test set using a probability of 0.5 as the classification threshold. The final classification model was then applied to the 186-sample validation set after data adjustment.

Processed data generated in this study are available within the article and its supplementary data files. Deidentified raw data generated in this study are available upon request from the corresponding author.

## Results

### Analyte Selection and Performance

Potential analytes were identified from prior studies that evaluated serum or plasma biomarkers in pre-malignant lesions, pre-diagnostic samples, or early-stage disease (Supplementary Table S2). Candidate biomarkers were selected for inclusion in the panel if identified in two or more independent experiments from blood products (stabilized whole blood, plasma, or serum) in early stage PDAC clinical samples. A subset of promising candidates that did not meet this standard was evaluated in pilot studies and candidate biomarkers were included for further analysis if the analyte was significantly elevated in early stage PDAC cases relative to healthy control subject (Supplementary Table S3). A total of 31 analytes were selected for inclusion in the biomarker panel (Table 2). Most of the analytes are secreted and/or function within the extracellular space and many participate in diverse biological processes associated with cancer including angiogenesis, inflammation, immune response, and cell migration (Supplementary Table S4).

**Table 2:**
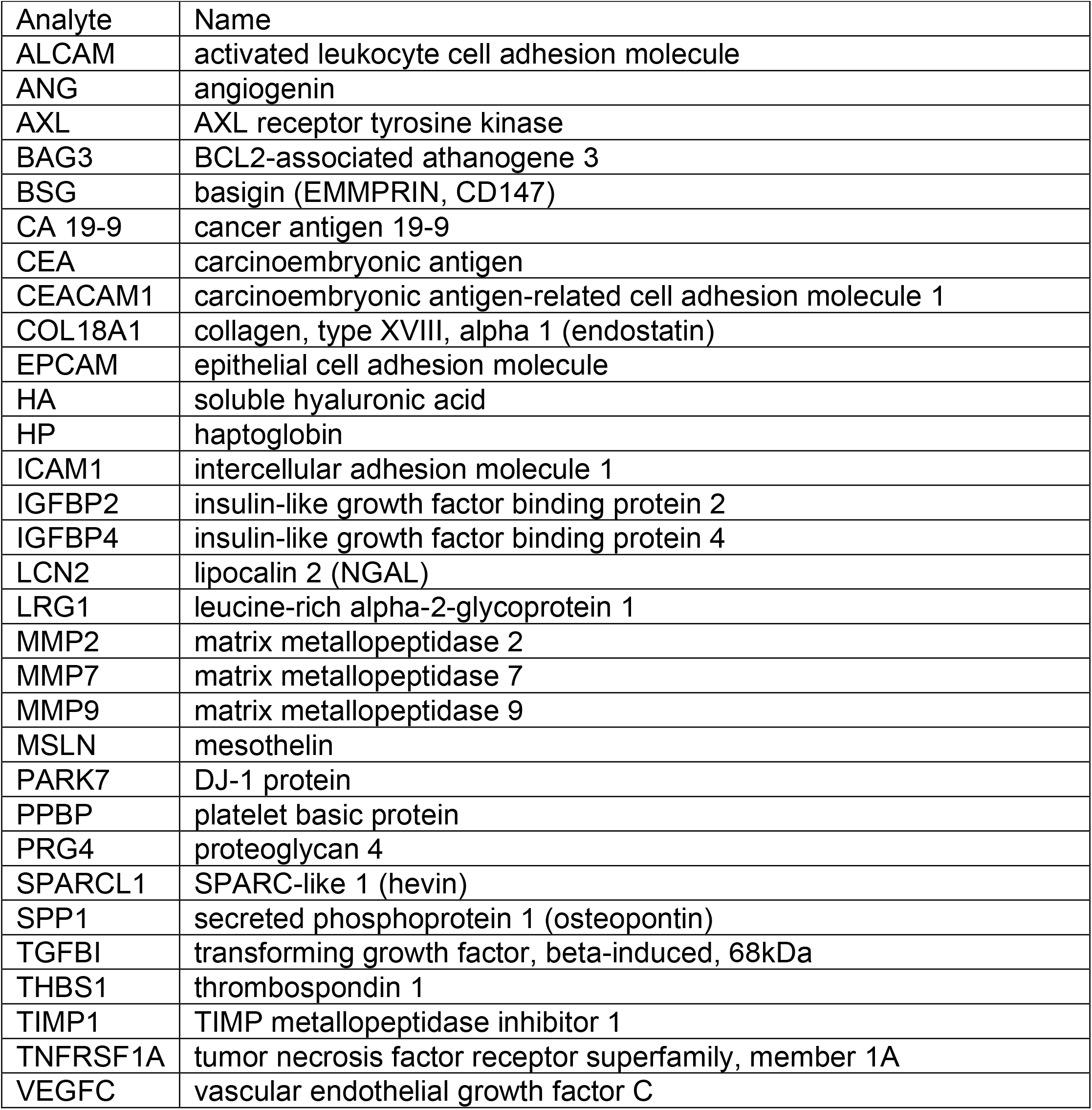
Analytes included in final panel

Levels of the selected analytes were measured in a development set of 837 serum samples from treatment-naïve patients with resectable PDAC, subjects with benign pancreatic disease, and subjects who were apparently healthy (Table 1). The diagnostic performance of the individual biomarkers was evaluated by ROC analyses (Supplementary Table S5). Twenty-nine of the 31 analytes yielded significantly higher area under the ROC curve than predicted by chance for at least one of the comparisons between PDAC and benign IPMN, chronic pancreatitis, or healthy controls demonstrating their potential for diagnostic discrimination of early stage PDAC. Two analytes, angiogenin (ANG) and matrix metallopeptidase 2 (MMP2) did not yield significant discrimination in this data set despite compelling evidence in the literature. Since the approach allows for interactions between analytes and because elevation of an individual analyte may occur in some PDAC cases, but not reach significance in aggregate data, ANG and MMP2 were included in the data sets for algorithm development, testing, and validation. Significant comparisons with increased AUC ranged from 0.53 to 0.84 with mean AUC of 0.64. CA 19-9 yielded the highest AUC for PDAC vs. healthy controls (AUC = 0.84) and chronic pancreatitis (AUC = 0.78), while CEACAM1 yielded the highest AUC for discrimination of PDAC from IPMN (AUC = 0.82). In general, the individual analytes were weak classifiers for discriminating PDAC from controls.

### Diagnostic Development and Performance

In an attempt to identify a stronger classifier, we used a machine learning approach to combine the information from the individual analytes (Figure 1). The approach proceeded in two steps. First, age, sex, and the 31 analyte levels in samples were provided as predictors to individual machine learning methods. The methods were chosen to represent dissimilar learning techniques (77) and pilot studies indicated that all predictors were utilized to some extent by one or more of the methods (Supplementary Table S6). The individual methods were used to assign two-class probabilities (early stage PDAC or non-PDAC control) for each case by optimizing ROC AUC via bootstrap re-sampling. Second, the outputs of the individual methods were combined by re-sampling into an ensemble model to assign the final class probabilities. The ensemble model was robust to sequential elimination of the individual methods, but each method contributed to model accuracy when specific combinations were considered (Supplementary Table S7).

**Figure 1:**
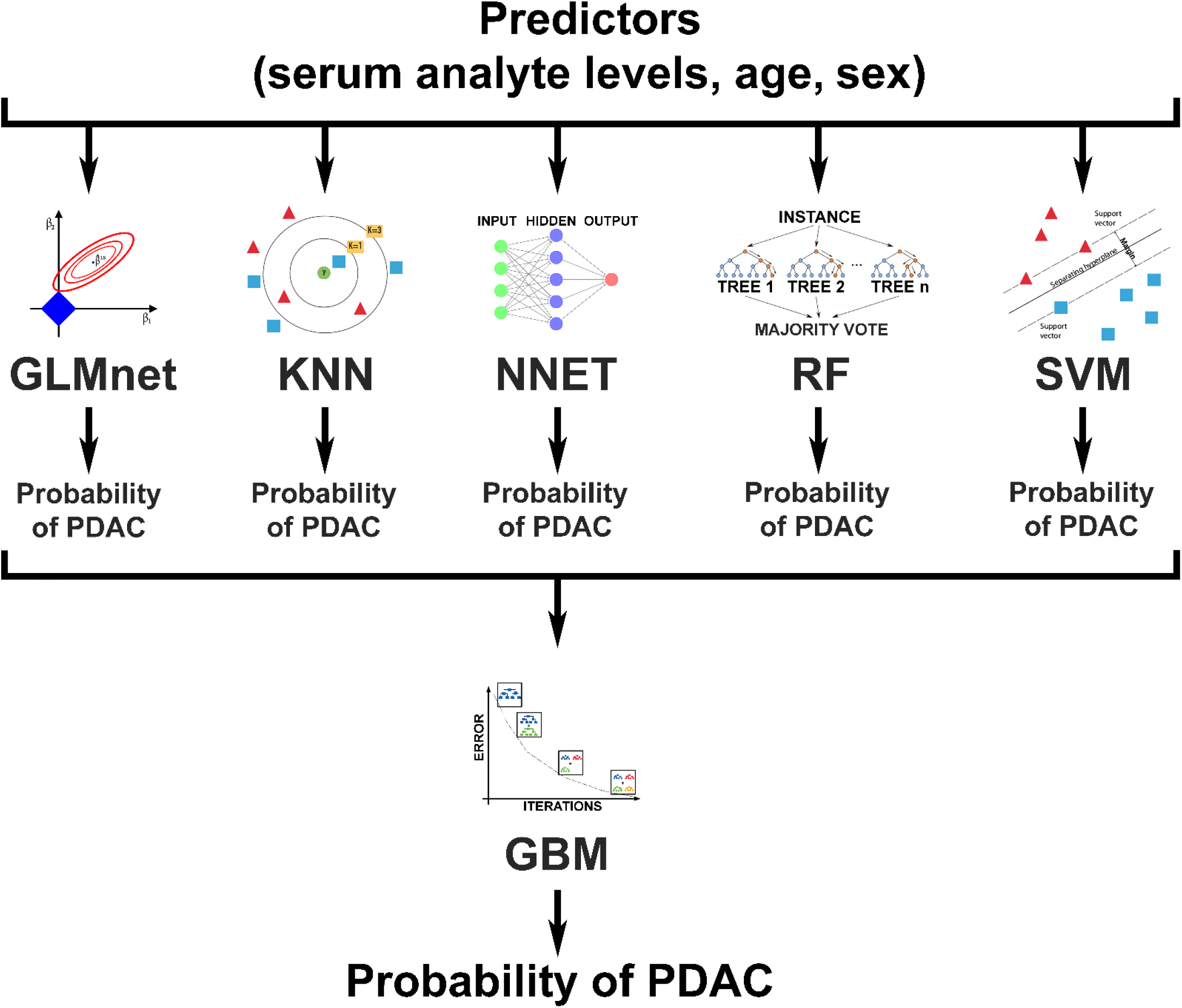
Machine learning schematic for diagnostic classification. Predictors were fed to individual machine learning methods and used to assign two-class probabilities (early stage PDAC or non-PDAC control) for each case. Outputs of the individual methods were combined into an ensemble model to assign the final class probabilities. GLMnet: Lasso and Elastic-Net Regularized Generalized Linear Models; KNN: k-Nearest Neighbors; NNET: Neural Network; RF: Random Forest; SVM: Support Vector Machines with Radial Basis Function Kernel; GBM: Generalized Boosted Regression Models.

The development set of 837 serum samples were randomly split into a training set representing 80% of the cases and a hold-out test set of 20% of cases. The two-step machine learning approach was applied to the training set to develop a diagnostic algorithm. The probabilities assigned by the algorithm to the training set resulted in a sensitivity of 92.8%, correctly identifying 141 of 152 PDAC cases. The algorithm yielded a specificity of 100%, correctly identifying all 358 healthy control cases. For benign pancreatic disease, the algorithm correctly identified all 82 chronic pancreatitis cases, but mis-identified 1 of 77 IPMN case for an overall specificity of 99.8% for identification of non-PDAC subjects (Table 3). The high specificity of the algorithm was a desirable result, given the rarity of the disease, but is likely a consequence of the higher ratio of controls to cases rather than a design feature, since overall diagnostic accuracy (case versus control) was used to optimize the algorithm.

**Table 3:** Diagnostic Performance

|  | Prediction |  |
| --- | --- | --- |
|  | PDAC | CON |
| <u>Training Set</u> |  |  |
| CON | 0 | 358 |
| ChPT | 0 | 82 |
| IPMN | 1 | 76 |
| PDAC | 141 | 11 |
| Accuracy | 0.982 |  |
| Sensitivity | 0.928 |  |
| Specificity |  |  |
| CON only | 1.0 |  |
| CON+IPMN+ChPT | 0.998 |  |
| <u>Test Set</u> |  |  |
| CON | 3 | 100 |
| ChPT | 2 | 16 |
| IPMN | 0 | 17 |
| PDAC | 19 | 11 |
| Accuracy | 0.905 |  |
| Sensitivity | 0.633 |  |
| Specificity |  |  |
| CON only | 0.971 |  |
| CON+IPMN+ChPT | 0.964 |  |
| <u>Validation Set</u> |  |  |
| CON | 2 | 38 |
| ChPT | 1 | 31 |
| Cystic Lesions (IPMN, MCN, SCA) | 2 | 39 |
| PDAC | 53 | 20 |
| Accuracy | 0.866 |  |
| Sensitivity | 0.726 |  |
| Specificity |  |  |
| CON only | 0.950 |  |
| CON+CL+ChPT | 0.956 |  |

For objective assessment of algorithm accuracy, the resulting algorithm was applied to unused samples from the hold-out test set resulting in a sensitivity of 63.3%, correctly identifying 19 of 30 PDAC cases and a specificity of 97.1% (100/101 healthy controls). All 17 IPMN cases were correctly identified, as were 16 of 18 chronic pancreatitis cases, yielding an overall specificity of 96.4% for identification of non-PDAC cases (Table 3). Receiver operating characteristic analysis illustrates that the diagnostic ability of the analyte panel for discrimination of PDAC from healthy controls (AUC = 0.944, Figure 2A) was greater than that of CA 19-9 alone (AUC = 0.878, Figure 2C) and for discrimination of PDAC from non-PDAC controls (analyte panel AUC = 0.920, Figure 2B vs. CA 19-9 AUC = 0.861, Figure 2D).

**Figure 2:**
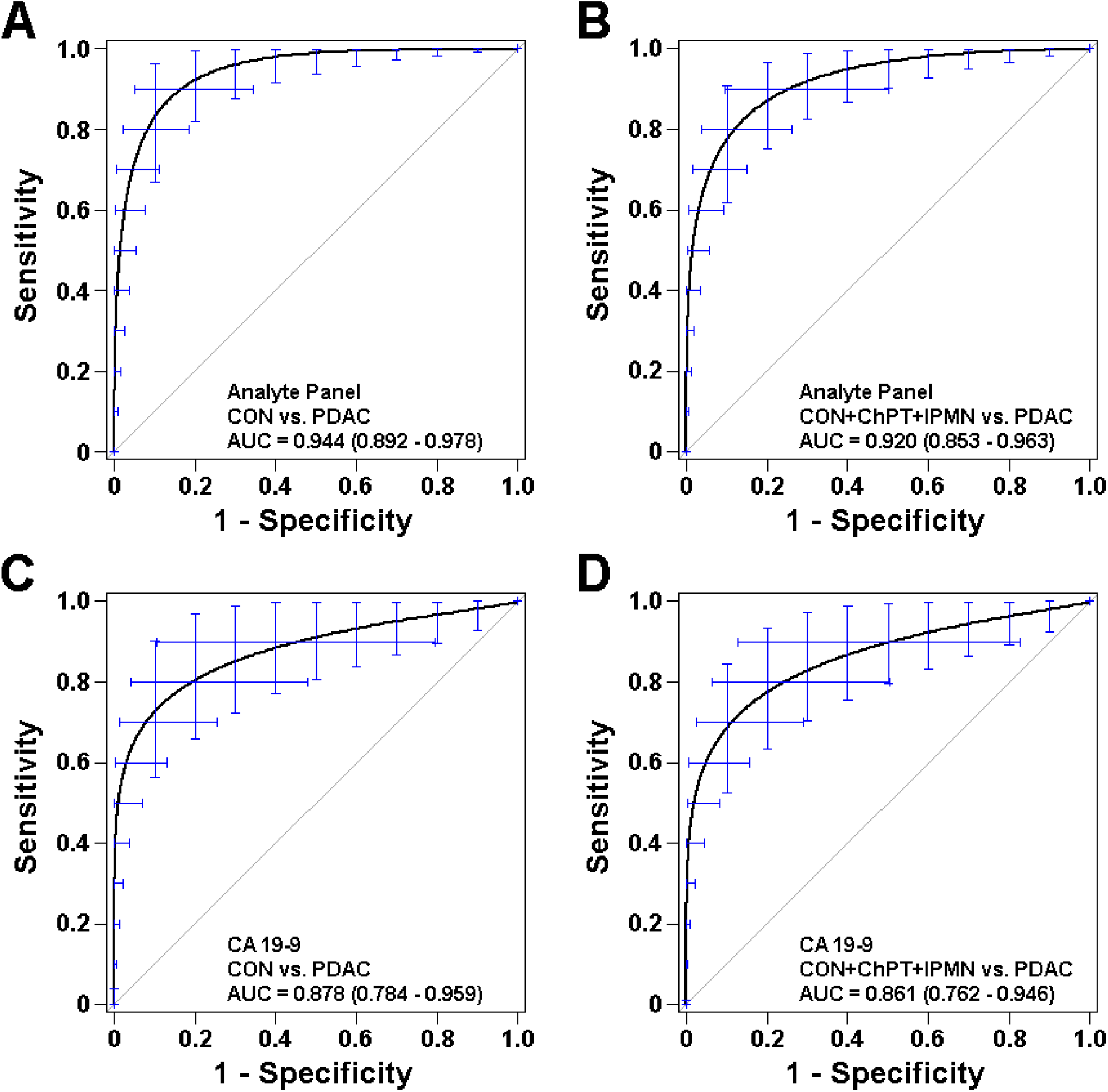
Diagnostic Performance in Test Set. Receiver operating characteristic curves describing the ability of the analyte panel for discrimination of early stage PDAC cases from (A) healthy controls and all non-PDAC controls including chronic pancreatitis, benign IPMN, and healthy controls (B) compared to similar discrimination by CA 19-9 alone (C, D) in the 20% hold-out test sample set. Bars represent 95% confidence intervals from 2000 bootstrap iterations.

### Diagnostic Validation

Additional samples, collected over a period of 18 months after the final analyte panel algorithm was constructed using the development set (Table 1), were used to evaluate the diagnostic utility of the algorithm in a real-world setting. The 31 analytes were measured in 186 additional serum samples representing 73 PDAC, 32 chronic pancreatitis, and 41 cystic lesions (27 IPMN, 5 mucinous cystic neoplasm, 9 serous cystadenoma) cases, as well as 40 samples from healthy control subjects. This data, when supplied to the analyte panel algorithm, correctly identifying 53 of the 73 PDAC cases resulting in a sensitivity of 72.6%, which was better performance than the test set. Classification of controls in the validation set was consistent with results in the test set with specificity of 95% (38/40) for healthy controls and 95.6% (108/113) for non-PDAC controls. In a rigorous validation using only those cases that satisfied the same inclusion criterion used for the development set (32 early stage PDAC, 32 chronic pancreatitis, 40 healthy controls [cystic lesions had less than the required 3 years follow-up]), the algorithm yielded a sensitivity of 81.5% (26/32) and specificity of 95% (38/40) for healthy controls and 95.8% (69/72) for non-PDAC controls.

For the validation set, all cases presenting with pancreatic disease were evaluated. While the development set only included early-stage resectable PDAC cases, the validation set included greater than 50% late-stage cases (Table 1), reflecting the expected distribution of stages at presentation. For cases with known stage, the analyte panel sensitivity was higher for stage I-II cases (81.3%) than for stage III-IV cases (63.9%), although the comparison was not significant (*P* = 0.27 by Fisher’s Exact Test). The development set also had the requirement that IPMN cases have at least 3 years of follow-up with no evidence of progression or high-grade dysplasia, which was not required for the validation set. Thus, cases with cystic lesions in the validation set may have included cases with more advanced disease, which cannot be excluded for the two cystic lesion cases in the validation set, both IPMN, that were classified as PDAC by the algorithm.

As with the test set, ROC analyses of the validation set showed greater diagnostic ability of the analyte panel for discrimination of PDAC than that of CA 19-9 alone. The analyte panel yielded an AUC of 0.925 for binary discrimination of PDAC from healthy controls (Figure 3A) compared to an AUC of 0.891 for CA 19-9 (Figure 3C). For discrimination of PDAC from all non-PDAC controls, the analyte panel had an AUC of 0.919 (Figure 3B) with CA 19-9 yielding an AUC of 0.874 (Figure 3D). For those PDAC cases that had CA 19-9 levels below the diagnostic threshold of 37 U/mL, the algorithm correctly classified 8 of 25 cases. These results confirm that the algorithm devised by machine learning in the development set was consistent in an independent sample set, show improvement over individual biomarkers, and suggest that the test may be useful for routine screening for early-stage disease in susceptible populations.

**Figure 3:**
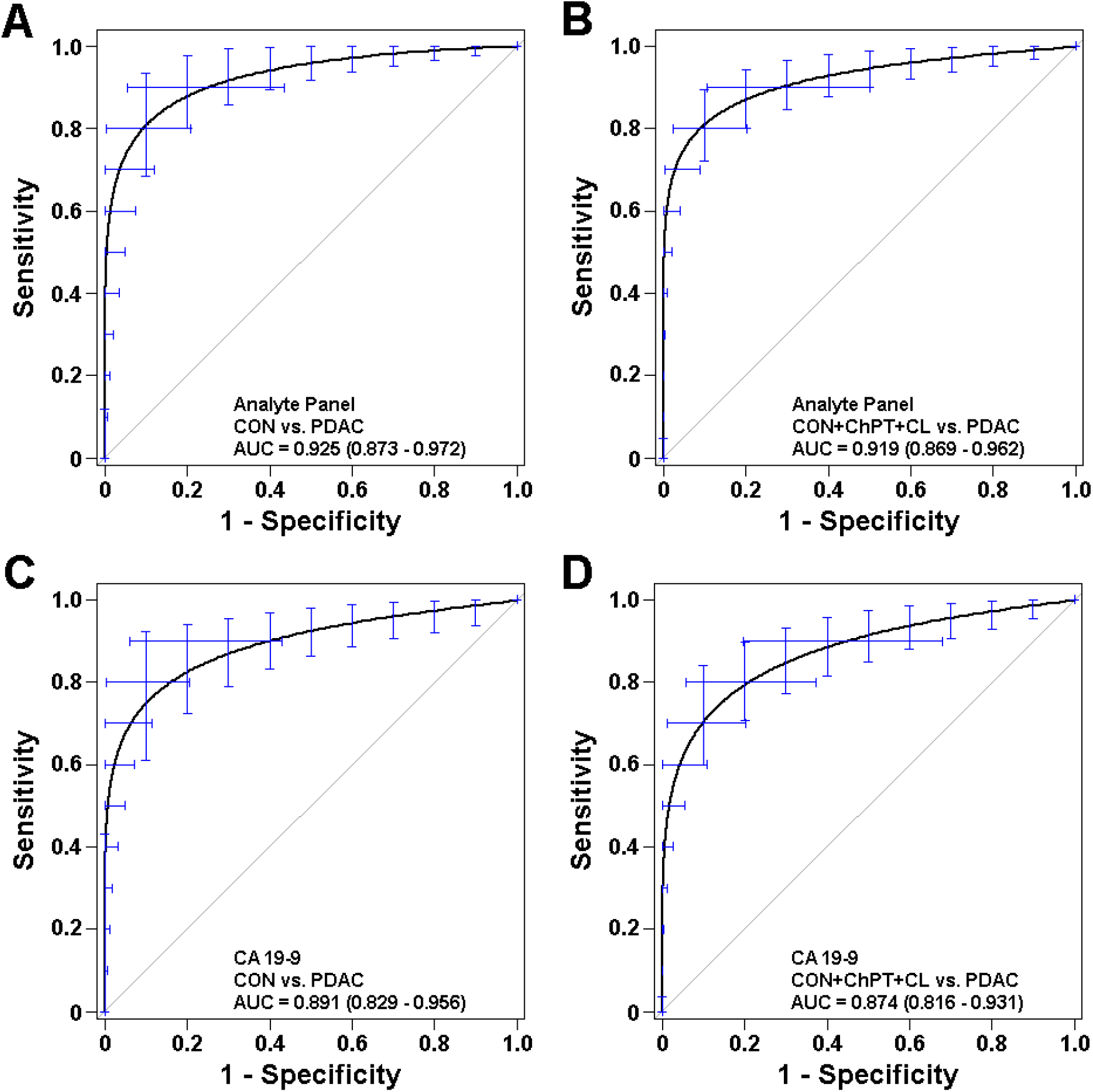
Diagnostic Performance in Validation Set. Receiver operating characteristic curves describing the ability of the analyte panel for discrimination of early stage PDAC cases from (A) healthy controls and all non-PDAC controls including chronic pancreatitis, cystic lesions (IPMN. MCN, SCA), and healthy controls (B) compared to similar discrimination by CA 19-9 alone (C, D) in the independent validation sample set. Bars represent 95% confidence intervals from 2000 bootstrap iterations.

## Discussion

Potential biomarkers evaluated for this study were previously identified as elevated in pre-malignant or early stage PDAC and largely validated in an extensive development sample set. The selected biomarkers participate in diverse biological processes, including angiogenesis, apoptosis, immune response, inflammatory response, and extracellular signal transduction. One barrier to diagnostic accuracy is that biomarker elevation may indicate unrelated conditions, but since these processes are also relevant to cancer, a large combined panel should be less ambiguous. A serial screening tool, deployed annually or biannually, would tend to obviate any unrelated acute condition that would be expected to resolve over the course of months unless it was also related to a chronic condition such as cancer. A second barrier to diagnostic accuracy is heterogeneity that may arise from biological, behavioral, and temporal differences between cases as well as controls. Attempts to find single or small sets of biomarkers would be more prone to the confounding of unrelated conditions and disease heterogeneity.

All predictors that met initial selection criterion were included throughout training, testing, and validation phases with the expectation that different subsets of biomarkers would more effectively discriminate cases and controls by accommodating disease heterogeneity (16,78). The fact that all predictors contributed to diagnostic models supports this expectation (Supplementary Table S6) and the differential utilization of predictors by the individual methods illustrate that different combinations of biomarkers contributed to discrimination. Two of the individual methods, KNN and SVM, had identical variable importance suggesting that these methods were redundant, but in method elimination analyses (Supplementary Table S7) in the ensemble model indicate that the two methods independently contribute to discrimination since elimination of SVM while keeping KNN reduced accuracy. This is likely due to machine learning optimization, rather than maximization. Derivation of the diagnostic algorithm was designed to be representative, not exhaustive, with the intent of minimizing over-fitting and devising an extensible algorithm.

The goal of machine learning is to reveal underlying structure of the data, in this case to assign binary probabilities for designation as either case or control classification. Selection of a single method optimized on one data set may emphasize set-specific or spurious aspects of the data organization that may not faithfully represent the data structure for a different data set. Ensemble methods seek to overcome the limitations of a single machine learning method by using resampling to evaluate performance of multiple machine learning models and then generate a combination of the models that optimizes test performance. This approach has been proven to be as accurate or better than the best individual component (79). A general risk for machine learning approaches is overfitting to a dataset that is unrepresentative to the general population; however, we found that our model gave almost identical AUC between test and validation, indicating that this algorithm may be generalizable. That the bootstrap accuracy of the test set, as measured by AUC, was 0.920 and the accuracy of the independent validation set was nearly identical (AUC = 0.919) suggests that the tradeoffs between under-computing and overfitting were reasonable. Consistent with expectation, the algorithm performance was greater than the best individual biomarker, CA 19-9, even though CA 19-9 performed very well in our sample sets and CA 19-9 assays achieved the highest range of historical accuracy (62).

Due to asymptomatic development, about 65% of PDAC cases present with late stage, unresectable disease (80). Biomarkers that increase with advancing stage and tumor burden are arguably specifically related to the presence of disease. To be effective in improving outcomes by shifting to lower stages at presentation, a PDAC diagnostic panel should be populated with analytes that are altered in early-stage disease and not just late-stage disease. The validation phase of this study included all cases presenting with pancreatic disease at our institute over the course of 18 months. For cases with known stage, the analyte panel sensitivity was higher for stage I-II cases (81.3%) than for stage III-IV cases (63.9%), although the comparison was not significant (*P* = 0.27 by Fisher’s Exact Test). These data suggest that tumor burden had limited contribution to the diagnostic determination and likely reflects prior selection for biomarkers specific for early-stage disease and algorithm training on only early-stage disease.

Screening programs for other, more common cancers, including breast, colorectal, and prostate cancers are now widespread with proven benefit; however, determining who receives the optimal benefit from these interventions is still debatable. Most screening programs run the risk of lead-time bias, overestimating the benefits of screening based on a perceived longer survival when the natural history of disease is not actually changed (81). Additionally, screening that identifies low-grade or premalignant neoplasms of unknown significance can lead to invasive testing and treatment with unknown clinical benefit (82,83). Any screening test must overcome these concerns.

Available evidence indicates that PDAC will almost always progress to metastases and ultimately death once malignant cells are present (84), but there appears to be a greater than 10-year lag time between first appearance of malignant cells and development of early-stage malignancy (11). A retrospective review of CT scans in PDAC patients obtained 12 – 18 months prior to diagnosis for unrelated issues demonstrates that subtle pancreatic abnormalities suggestive of PDAC are often seen (85). While this demonstrates a window for early detection, it also shows the real concern for lead time bias. Ultimately, proof of efficacy for any screening tool for PDAC will require prospective evaluation in a larger population demonstrating not only improved survival, but also decreased mortality. Also, as this study was largely a single institution investigation the results will require validation in a multi-institutional cohort to increase external validity.

Any screening test for PDAC must be highly specific to avoid false-positive screens leading to increased morbidity, cost, and emotional distress associated with secondary screening tests and treatment (86). The recommendation that screening be limited to high-risk groups (87) would increase pre-test probability and reduce the number of false-positive diagnoses, as PDAC has a far lower prevalence in the general population than other commonly screened malignancies. Screening with imaging is relatively low risk, although endoscopy is associated with low rates of post-procedural pain, anesthesia related adverse events, and acute pancreatitis (88,89). Screening with primary imaging is expensive however, with one study estimating the cost to detect a single PDAC in a high-risk cohort is over $41,000 (90). The analytes and algorithms devised in this study may be useful as a primary screening tool for assessing the need for secondary screening by imaging while limiting concerns of unnecessary morbidity caused by false positive determinations due to its high specificity. The analyses routinely yielded the desirable high specificity, although with lower, but actionable sensitivity. In a serial screening program, the test will have the opportunity to increase overall sensitivity by re-evaluation of negative initial tests.

The analyses show that the general approach to PDAC screening is valid, that our samples generate reproducible signals, and that novel analytes contribute to disease classification. This 31-biomarker assay may be helpful in developing a useful tool for identifying early-stage, asymptomatic PDAC, particularly for screening in high-risk patient populations to guide the use of screening imaging. The results also serve to illustrate the proposed analytical methods which allows for interaction between analytes, subsets, and heterogenous biological response amongst subjects. Finally, the approach allows for improvement through incorporation of additional biomarkers, including signals from cell-free DNA, circulating tumor cells, and demographic factors.

## Data Availability

All data produced in the present study are available upon reasonable request to the authors.

## Notes

This study was partially supported by research grants from the National Institutes of Health (CA115225, CA151650, CA155586, CA196403, CA200468 to SJM and P30CA042014 to the Huntsman Cancer Institute for support of core facilities), grants from the Huntsman Cancer Institute Gastrointestinal Cancer Research Program and through support from the Huntsman Cancer Foundation (MAF, SJM).

MAF and SJM are inventors named on a patent held by the University of Utah for claims associated with this project. MCT, AR, VDL, LM, MDM and AF are shareholders of BIOUNIVERSA s.r.l. that is involved in the development of BAG3 specific diagnostic reagents. MCT is inventor of a patent in the field of BAG3 used as a biomarker in the diagnosis of pancreatic cancer. The other authors declare no potential conflicts of interest.

### Funding Statement

This study was partially supported by research grants from the National Institutes of Health to SJM and to the Huntsman Cancer Institute for support of core facilities, grants from the Huntsman Cancer Institute Gastrointestinal Cancer Research Program and through support from the Huntsman Cancer Foundation.

### Author Declarations

The IRB of the University of Utah gave ethical approval for this work.

## REFERENCES

1. The Global Cancer Observatory G. Source: Globocan 2018. World Heal Organ. 2019;876:2018–2019. <https://gco.iarc.fr>.

2. Siegel RL, Miller KD, Jemal A. Cancer statistics, 2020. CA: a cancer journal for clinicians 2020;70(1):7–30 doi 10.3322/caac.21590.

3. Saad AM, Turk T, Al-Husseini MJ, Abdel-Rahman O. Trends in pancreatic adenocarcinoma incidence and mortality in the United States in the last four decades; a SEER-based study. BMC cancer 2018;18(1):688 doi 10.1186/s12885-018-4610-4.

4. Rahib L, Smith BD, Aizenberg R, Rosenzweig AB, Fleshman JM, Matrisian LM. Projecting cancer incidence and deaths to 2030: the unexpected burden of thyroid, liver, and pancreas cancers in the United States. Cancer Res 2014;74(11):2913–21 doi 10.1158/0008-5472.CAN-14-0155.

5. Vincent A, Herman J, Schulick R, Hruban RH, Goggins M. Pancreatic cancer. Lancet 2011;378(9791):607–20 doi 10.1016/S0140-6736(10)62307-0.

6. Acher AW, Bleicher J, Cannon A, Scaife C. Advances in surgery for pancreatic cancer. Journal of gastrointestinal oncology 2018;9(6):1037–43 doi 10.21037/jgo.2018.05.05.

7. Bockhorn M, Uzunoglu FG, Adham M, Imrie C, Milicevic M, Sandberg AA, et al. Borderline resectable pancreatic cancer: a consensus statement by the International Study Group of Pancreatic Surgery (ISGPS). Surgery 2014;155(6):977–88 doi 10.1016/j.surg.2014.02.001.

8. Heinrich S, Besselink M, Moehler M, van Laethem JL, Ducreux M, Grimminger P, et al. Opinions and use of neoadjuvant therapy for resectable, borderline resectable, and locally advanced pancreatic cancer: international survey and case-vignette study. BMC cancer 2019;19(1):675 doi 10.1186/s12885-019-5889-5.

9. Ilic M, Ilic I. Epidemiology of pancreatic cancer. World J Gastroenterol 2016;22(44):9694–705 doi 10.3748/wjg.v22.i44.9694.

10. National Cancer Institute - Surveillance Epidemiology and End Results Program. Cancer Stat Facts: Pancreatic Cancer. <https://seer.cancer.gov/statfacts/html/pancreas.html>.

11. Yachida S, Jones S, Bozic I, Antal T, Leary R, Fu B, et al. Distant metastasis occurs late during the genetic evolution of pancreatic cancer. Nature 2010;467(7319):1114–7 doi 10.1038/nature09515.

12. Ryan DP, Hong TS, Bardeesy N. Pancreatic adenocarcinoma. The New England journal of medicine 2014;371(22):2140–1 doi 10.1056/NEJMc1412266.

13. Owens DK, Davidson KW, Krist AH, Barry MJ, Cabana M, Caughey AB, et al. Screening for Pancreatic Cancer: US Preventive Services Task Force Reaffirmation Recommendation Statement. Jama 2019;322(5):438–44 doi 10.1001/jama.2019.10232.

14. Rubenstein JH, Scheiman JM, Anderson MA. A clinical and economic evaluation of endoscopic ultrasound for patients at risk for familial pancreatic adenocarcinoma. Pancreatology : official journal of the International Association of Pancreatology 2007;7(5-6):514–25 doi 10.1159/000108969.

15. Rulyak SJ, Kimmey MB, Veenstra DL, Brentnall TA. Cost-effectiveness of pancreatic cancer screening in familial pancreatic cancer kindreds. Gastrointestinal endoscopy 2003;57(1):23–9 doi 10.1067/mge.2003.28.

16. Firpo MA, Boucher KM, Mulvihill SJ. Prospects for developing an accurate diagnostic biomarker panel for low prevalence cancers. Theoretical biology & medical modelling 2014;11:34 doi 10.1186/1742-4682-11-34.

17. Kim J, Bamlet WR, Oberg AL, Chaffee KG, Donahue G, Cao XJ, et al. Detection of early pancreatic ductal adenocarcinoma with thrombospondin-2 and CA19-9 blood markers. Science translational medicine 2017;9(398) doi 10.1126/scitranslmed.aah5583.

18. Argani P, Iacobuzio-Donahue C, Ryu B, Rosty C, Goggins M, Wilentz RE, et al. Mesothelin is overexpressed in the vast majority of ductal adenocarcinomas of the pancreas: identification of a new pancreatic cancer marker by serial analysis of gene expression (SAGE). Clinical cancer research : an official journal of the American Association for Cancer Research 2001;7(12):3862–8.

19. Bramhall SR, Neoptolemos JP, Stamp GW, Lemoine NR. Imbalance of expression of matrix metalloproteinases (MMPs) and tissue inhibitors of the matrix metalloproteinases (TIMPs) in human pancreatic carcinoma. J Pathol 1997;182(3):347–55 doi 10.1002/(SICI)1096-9896(199707)182:3<347::AID-PATH848>3.0.CO;2-J.

20. Buchholz M, Braun M, Heidenblut A, Kestler HA, Kloppel G, Schmiegel W, et al. Transcriptome analysis of microdissected pancreatic intraepithelial neoplastic lesions. Oncogene 2005;24(44):6626–36 doi 10.1038/sj.onc.1208804.

21. Chen R, Crispin DA, Pan S, Hawley S, McIntosh MW, May D, et al. Pilot study of blood biomarker candidates for detection of pancreatic cancer. Pancreas 2010;39(7):981–8 doi 10.1097/MPA.0b013e3181dac920.

22. Chen R, Yi EC, Donohoe S, Pan S, Eng J, Cooke K, et al. Pancreatic cancer proteome: the proteins that underlie invasion, metastasis, and immunologic escape. Gastroenterology 2005;129(4):1187–97 doi 10.1053/j.gastro.2005.08.001.

23. Coppola D, Szabo M, Boulware D, Muraca P, Alsarraj M, Chambers AF, et al. Correlation of osteopontin protein expression and pathological stage across a wide variety of tumor histologies. Clinical cancer research : an official journal of the American Association for Cancer Research 2004;10(1 Pt 1):184–90 doi 10.1158/1078-0432.ccr-1405-2.

24. Del Villano BC, Brennan S, Brock P, Bucher C, Liu V, McClure M, et al. Radioimmunometric assay for a monoclonal antibody-defined tumor marker, CA 19-9. Clinical chemistry 1983;29(3):549–52.

25. Ellenrieder V, Alber B, Lacher U, Hendler SF, Menke A, Boeck W, et al. Role of MT-MMPs and MMP-2 in pancreatic cancer progression. International journal of cancer 2000;85(1):14–20 doi 10.1002/(sici)1097-0215(20000101)85:1<14::aid-ijc3>3.0.co;2-o.

26. Esposito I, Kayed H, Keleg S, Giese T, Sage EH, Schirmacher P, et al. Tumor-suppressor function of SPARC-like protein 1/Hevin in pancreatic cancer. Neoplasia 2007;9(1):8–17 doi 10.1593/neo.06646.

27. Faca VM, Song KS, Wang H, Zhang Q, Krasnoselsky AL, Newcomb LF, et al. A mouse to human search for plasma proteome changes associated with pancreatic tumor development. PLoS medicine 2008;5(6):e123 doi 10.1371/journal.pmed.0050123.

28. Falco A, Rosati A, Festa M, Basile A, De Marco M, d’Avenia M, et al. BAG3 is a novel serum biomarker for pancreatic adenocarcinomas. The American journal of gastroenterology 2013;108(7):1178–80 doi 10.1038/ajg.2013.128.

29. Firpo MA, Gay DZ, Granger SR, Scaife CL, DiSario JA, Boucher KM, et al. Improved diagnosis of pancreatic adenocarcinoma using haptoglobin and serum amyloid A in a panel screen. World journal of surgery 2009;33(4):716–22 doi 10.1007/s00268-008-9853-9.

30. Gebauer F, Struck L, Tachezy M, Vashist Y, Wicklein D, Schumacher U, et al. Serum EpCAM expression in pancreatic cancer. Anticancer Res 2014;34(9):4741–6.

31. Giusti G, Piccinino F, Sagnelli E, Pasquale G. Differential diagnosis between benign and malignant biliary tract obstruction: discriminative usefulness of some clinical and laboratory data. Acta hepatogastroenterologica 1975;22(6):374–9.

32. Grutzmann R, Pilarsky C, Ammerpohl O, Luttges J, Bohme A, Sipos B, et al. Gene expression profiling of microdissected pancreatic ductal carcinomas using high-density DNA microarrays. Neoplasia 2004;6(5):611–22 doi 10.1593/neo.04295.

33. Harsha HC, Kandasamy K, Ranganathan P, Rani S, Ramabadran S, Gollapudi S, et al. A compendium of potential biomarkers of pancreatic cancer. PLoS medicine 2009;6(4):e1000046 doi 10.1371/journal.pmed.1000046.

34. Harvey SR, Hurd TC, Markus G, Martinick MI, Penetrante RM, Tan D, et al. Evaluation of urinary plasminogen activator, its receptor, matrix metalloproteinase-9, and von Willebrand factor in pancreatic cancer. Clinical cancer research : an official journal of the American Association for Cancer Research 2003;9(13):4935–43.

35. Hassan R, Laszik ZG, Lerner M, Raffeld M, Postier R, Brackett D. Mesothelin is overexpressed in pancreaticobiliary adenocarcinomas but not in normal pancreas and chronic pancreatitis. Am J Clin Pathol 2005;124(6):838–45.

36. He XY, Liu BY, Yao WY, Zhao XJ, Zheng Z, Li JF, et al. Serum DJ-1 as a diagnostic marker and prognostic factor for pancreatic cancer. Journal of digestive diseases 2011;12(2):131–7 doi 10.1111/j.1751-2980.2011.00488.x.

37. Holyoke D, Reynoso G, Chu TM. Carcinoembryonic antigen (CEA) in patients with carcinoma of the digestive tract. Annals of surgery 1972;176(4):559–64 doi 10.1097/00000658-197217640-00015.

38. Iacobuzio-Donahue CA, Maitra A, Olsen M, Lowe AW, van Heek NT, Rosty C, et al. Exploration of global gene expression patterns in pancreatic adenocarcinoma using cDNA microarrays. Am J Pathol 2003;162(4):1151–62 doi 10.1016/S0002-9440(10)63911-9.

39. Iacobuzio-Donahue CA, Ryu B, Hruban RH, Kern SE. Exploring the host desmoplastic response to pancreatic carcinoma: gene expression of stromal and neoplastic cells at the site of primary invasion. Am J Pathol 2002;160(1):91–9 doi 10.1016/S0002-9440(10)64353-2.

40. Johnston FM, Tan MC, Tan BR, Jr., Porembka MR, Brunt EM, Linehan DC, et al. Circulating mesothelin protein and cellular antimesothelin immunity in patients with pancreatic cancer. Clinical cancer research : an official journal of the American Association for Cancer Research 2009;15(21):6511–8 doi 10.1158/1078-0432.CCR-09-0565.

41. Kakisaka T, Kondo T, Okano T, Fujii K, Honda K, Endo M, et al. Plasma proteomics of pancreatic cancer patients by multi-dimensional liquid chromatography and two-dimensional difference gel electrophoresis (2D-DIGE): up-regulation of leucine-rich alpha-2-glycoprotein in pancreatic cancer. Journal of chromatography B, Analytical technologies in the biomedical and life sciences 2007;852(1-2):257–67 doi 10.1016/j.jchromb.2007.01.029.

42. Kasper HU, Ebert M, Malfertheiner P, Roessner A, Kirkpatrick CJ, Wolf HK. Expression of thrombospondin-1 in pancreatic carcinoma: correlation with microvessel density. Virchows Arch 2001;438(2):116–20 doi 10.1007/s004280000302.

43. Kendrick ZW, Firpo MA, Repko RC, Scaife CL, Adler DG, Boucher KM, et al. Serum IGFBP2 and MSLN as diagnostic and prognostic biomarkers for pancreatic cancer. HPB : the official journal of the International Hepato Pancreato Biliary Association 2014;16(7):670–6 doi 10.1111/hpb.12199.

44. Kolb A, Kleeff J, Guweidhi A, Esposito I, Giese NA, Adwan H, et al. Osteopontin influences the invasiveness of pancreatic cancer cells and is increased in neoplastic and inflammatory conditions. Cancer biology & therapy 2005;4(7):740–6 doi 10.4161/cbt.4.7.1821.

45. Koopmann J, Fedarko NS, Jain A, Maitra A, Iacobuzio-Donahue C, Rahman A, et al. Evaluation of osteopontin as biomarker for pancreatic adenocarcinoma. Cancer epidemiology, biomarkers & prevention : a publication of the American Association for Cancer Research, cosponsored by the American Society of Preventive Oncology 2004;13(3):487–91.

46. Koopmann J, Rosenzweig CN, Zhang Z, Canto MI, Brown DA, Hunter M, et al. Serum markers in patients with resectable pancreatic adenocarcinoma: macrophage inhibitory cytokine 1 versus CA19-9. Clinical cancer research : an official journal of the American Association for Cancer Research 2006;12(2):442–6 doi 10.1158/1078-0432.CCR-05-0564.

47. Kuhlmann KF, van Till JW, Boermeester MA, de Reuver PR, Tzvetanova ID, Offerhaus GJ, et al. Evaluation of matrix metalloproteinase 7 in plasma and pancreatic juice as a biomarker for pancreatic cancer. Cancer epidemiology, biomarkers & prevention : a publication of the American Association for Cancer Research, cosponsored by the American Society of Preventive Oncology 2007;16(5):886–91 doi 10.1158/1055-9965.EPI-06-0779.

48. Laurell H, Bouisson M, Berthelemy P, Rochaix P, Dejean S, Besse P, et al. Identification of biomarkers of human pancreatic adenocarcinomas by expression profiling and validation with gene expression analysis in endoscopic ultrasound-guided fine needle aspiration samples. World J Gastroenterol 2006;12(21):3344–51 doi 10.3748/wjg.v12.i21.3344.

49. Li M, Zhai Q, Bharadwaj U, Wang H, Li F, Fisher WE, et al. Cyclophilin A is overexpressed in human pancreatic cancer cells and stimulates cell proliferation through CD147. Cancer 2006;106(10):2284–94 doi 10.1002/cncr.21862.

50. Li YJ, Wei ZM, Meng YX, Ji XR. Beta-catenin up-regulates the expression of cyclinD1, c-myc and MMP-7 in human pancreatic cancer: relationships with carcinogenesis and metastasis. World J Gastroenterol 2005;11(14):2117–23 doi 10.3748/wjg.v11.i14.2117.

51. Liao Q, Ozawa F, Friess H, Zimmermann A, Takayama S, Reed JC, et al. The anti-apoptotic protein BAG-3 is overexpressed in pancreatic cancer and induced by heat stress in pancreatic cancer cell lines. FEBS Lett 2001;503(2-3):151–7 doi 10.1016/s0014-5793(01)02728-4.

52. Logsdon CD, Simeone DM, Binkley C, Arumugam T, Greenson JK, Giordano TJ, et al. Molecular profiling of pancreatic adenocarcinoma and chronic pancreatitis identifies multiple genes differentially regulated in pancreatic cancer. Cancer Res 2003;63(10):2649–57.

53. Mahlbacher V, Sewing A, Elsasser HP, Kern HF. Hyaluronan is a secretory product of human pancreatic adenocarcinoma cells. Eur J Cell Biol 1992;58(1):28–34.

54. Maitra A, Adsay NV, Argani P, Iacobuzio-Donahue C, De Marzo A, Cameron JL, et al. Multicomponent analysis of the pancreatic adenocarcinoma progression model using a pancreatic intraepithelial neoplasia tissue microarray. Mod Pathol 2003;16(9):902–12 doi 10.1097/01.MP.0000086072.56290.FB.

55. Melle C, Ernst G, Escher N, Hartmann D, Schimmel B, Bleul A, et al. Protein profiling of microdissected pancreas carcinoma and identification of HSP27 as a potential serum marker. Clinical chemistry 2007;53(4):629–35 doi 10.1373/clinchem.2006.079194.

56. Missiaglia E, Blaveri E, Terris B, Wang YH, Costello E, Neoptolemos JP, et al. Analysis of gene expression in cancer cell lines identifies candidate markers for pancreatic tumorigenesis and metastasis. International journal of cancer 2004;112(1):100–12 doi 10.1002/ijc.20376.

57. Ohlund D, Ardnor B, Oman M, Naredi P, Sund M. Expression pattern and circulating levels of endostatin in patients with pancreas cancer. International journal of cancer 2008;122(12):2805–10 doi 10.1002/ijc.23468.

58. Ordonez NG. Application of mesothelin immunostaining in tumor diagnosis. Am J Surg Pathol 2003;27(11):1418–28 doi 10.1097/00000478-200311000-00003.

59. Pan S, Chen R, Crispin DA, May D, Stevens T, McIntosh MW, et al. Protein alterations associated with pancreatic cancer and chronic pancreatitis found in human plasma using global quantitative proteomics profiling. Journal of proteome research 2011;10(5):2359–76 doi 10.1021/pr101148r.

60. Park HD, Kang ES, Kim JW, Lee KT, Lee KH, Park YS, et al. Serum CA19-9, cathepsin D, and matrix metalloproteinase-7 as a diagnostic panel for pancreatic ductal adenocarcinoma. Proteomics 2012;12(23-24):3590–7 doi 10.1002/pmic.201200101.

61. Poruk KE, Firpo MA, Scaife CL, Adler DG, Emerson LL, Boucher KM, et al. Serum osteopontin and tissue inhibitor of metalloproteinase 1 as diagnostic and prognostic biomarkers for pancreatic adenocarcinoma. Pancreas 2013;42(2):193–7 doi 10.1097/MPA.0b013e31825e354d.

62. Poruk KE, Gay DZ, Brown K, Mulvihill JD, Boucher KM, Scaife CL, et al. The clinical utility of CA 19-9 in pancreatic adenocarcinoma: diagnostic and prognostic updates. Current molecular medicine 2013;13(3):340–51 doi 10.2174/1566524011313030003.

63. Qian X, Rothman VL, Nicosia RF, Tuszynski GP. Expression of thrombospondin-1 in human pancreatic adenocarcinomas: role in matrix metalloproteinase-9 production. Pathol Oncol Res 2001;7(4):251–9 doi 10.1007/bf03032381.

64. Riethdorf S, Reimers N, Assmann V, Kornfeld JW, Terracciano L, Sauter G, et al. High incidence of EMMPRIN expression in human tumors. International journal of cancer 2006;119(8):1800–10 doi 10.1002/ijc.22062.

65. Schneiderhan W, Diaz F, Fundel M, Zhou S, Siech M, Hasel C, et al. Pancreatic stellate cells are an important source of MMP-2 in human pancreatic cancer and accelerate tumor progression in a murine xenograft model and CAM assay. J Cell Sci 2007;120(Pt 3):512–9 doi 10.1242/jcs.03347.

66. Shimoyama S, Gansauge F, Gansauge S, Negri G, Oohara T, Beger HG. Increased angiogenin expression in pancreatic cancer is related to cancer aggressiveness. Cancer Res 1996;56(12):2703–6.

67. Shimoyama S, Gansauge F, Gansauge S, Oohara T, Kaminishi M, Beger HG. Increased angiogenin expression in obstructive chronic pancreatitis surrounding pancreatic cancer but not in pure chronic pancreatitis. Pancreas 1999;18(3):225–30 doi 10.1097/00006676-199904000-00002.

68. Simeone DM, Ji B, Banerjee M, Arumugam T, Li D, Anderson MA, et al. CEACAM1, a novel serum biomarker for pancreatic cancer. Pancreas 2007;34(4):436–43 doi 10.1097/MPA.0b013e3180333ae3.

69. Tang RF, Itakura J, Aikawa T, Matsuda K, Fujii H, Korc M, et al. Overexpression of lymphangiogenic growth factor VEGF-C in human pancreatic cancer. Pancreas 2001;22(3):285–92 doi 10.1097/00006676-200104000-00010.

70. Xie MJ, Motoo Y, Su SB, Mouri H, Ohtsubo K, Matsubara F, et al. Expression of clusterin in human pancreatic cancer. Pancreas 2002;25(3):234–8 doi 10.1097/00006676-200210000-00004.

71. Zeh HJ, Winikoff S, Landsittel DP, Gorelik E, Marrangoni AM, Velikokhatnaya L, et al. Multianalyte profiling of serum cytokines for detection of pancreatic cancer. Cancer Biomark 2005;1(6):259–69 doi 10.3233/cbm-2005-1601.

72. Zhang W, Erkan M, Abiatari I, Giese NA, Felix K, Kayed H, et al. Expression of extracellular matrix metalloproteinase inducer (EMMPRIN/CD147) in pancreatic neoplasm and pancreatic stellate cells. Cancer biology & therapy 2007;6(2):218–27 doi 10.4161/cbt.6.2.3623.

73. Zhou W, Sokoll LJ, Bruzek DJ, Zhang L, Velculescu VE, Goldin SB, et al. Identifying markers for pancreatic cancer by gene expression analysis. Cancer epidemiology, biomarkers & prevention : a publication of the American Association for Cancer Research, cosponsored by the American Society of Preventive Oncology 1998;7(2):109–12.

74. Basile A, De Marco M, Festa M, Falco A, Iorio V, Guerriero L, et al. Development of an anti-BAG3 humanized antibody for treatment of pancreatic cancer. Molecular oncology 2019;13(6):1388–99 doi 10.1002/1878-0261.12492.

75. R Core Team. 2019 R: A language and environment for statistical computing. R Foundation for Statistical Computing, Vienna, Austria. <https://www.R-project.org/>.

76. Deane-Mayer ZA, Knowles JE. 2016 caretEnsemble: Ensembles of Caret Models. R package version 2.0.0. <https://CRAN.R-project.org/package=caretEnsemble>.

77. Kuhn M. The caret Package. <https://topepo.github.io/caret/models-clustered-by-tag-similarity.html>.

78. Crawford HC, Wallace MB, Storz P. Early detection and imaging strategies to reveal and target developing pancreatic cancer. Expert Rev Anticancer Ther 2020;20(2):81–3 doi 10.1080/14737140.2020.1720654.

79. van der Laan MJ, Polley EC, Hubbard AE. Super learner. Stat Appl Genet Mol Biol 2007;6:Article25 doi 10.2202/1544-6115.1309.

80. Chari ST, Kelly K, Hollingsworth MA, Thayer SP, Ahlquist DA, Andersen DK, et al. Early detection of sporadic pancreatic cancer: summative review. Pancreas 2015;44(5):693–712 doi 10.1097/MPA.0000000000000368.

81. Poruk KE, Firpo MA, Mulvihill SJ. Screening for pancreatic cancer. Advances in surgery 2014;48:115–36 doi 10.1016/j.yasu.2014.05.004.

82. Schwartz LM, Woloshin S, Fowler FJ, Jr., Welch HG. Enthusiasm for cancer screening in the United States. Jama 2004;291(1):71–8 doi 10.1001/jama.291.1.71.

83. Welch HG, Fisher ES. Income and Cancer Overdiagnosis - When Too Much Care Is Harmful. The New England journal of medicine 2017;376(23):2208–9 doi 10.1056/NEJMp1615069.

84. McGuigan A, Kelly P, Turkington RC, Jones C, Coleman HG, McCain RS. Pancreatic cancer: A review of clinical diagnosis, epidemiology, treatment and outcomes. World J Gastroenterol 2018;24(43):4846–61 doi 10.3748/wjg.v24.i43.4846.

85. Pelaez-Luna M, Takahashi N, Fletcher JG, Chari ST. Resectability of presymptomatic pancreatic cancer and its relationship to onset of diabetes: a retrospective review of CT scans and fasting glucose values prior to diagnosis. The American journal of gastroenterology 2007;102(10):2157–63 doi 10.1111/j.1572-0241.2007.01480.x.

86. Poruk KE, Firpo MA, Adler DG, Mulvihill SJ. Screening for pancreatic cancer: why, how, and who? Annals of surgery 2013;257(1):17–26 doi 10.1097/SLA.0b013e31825ffbfb.

87. Canto MI, Harinck F, Hruban RH, Offerhaus GJ, Poley JW, Kamel I, et al. International Cancer of the Pancreas Screening (CAPS) Consortium summit on the management of patients with increased risk for familial pancreatic cancer. Gut 2013;62(3):339–47 doi 10.1136/gutjnl-2012-303108.

88. Canto MI, Goggins M, Hruban RH, Petersen GM, Giardiello FM, Yeo C, et al. Screening for early pancreatic neoplasia in high-risk individuals: a prospective controlled study. Clinical gastroenterology and hepatology : the official clinical practice journal of the American Gastroenterological Association 2006;4(6):766–81; quiz 665 doi 10.1016/j.cgh.2006.02.005.

89. Canto MI, Goggins M, Yeo CJ, Griffin C, Axilbund JE, Brune K, et al. Screening for pancreatic neoplasia in high-risk individuals: an EUS-based approach. Clinical gastroenterology and hepatology : the official clinical practice journal of the American Gastroenterological Association 2004;2(7):606–21 doi 10.1016/s1542-3565(04)00244-7.

90. Zubarik R, Gordon SR, Lidofsky SD, Anderson SR, Pipas JM, Badger G, et al. Screening for pancreatic cancer in a high-risk population with serum CA 19-9 and targeted EUS: a feasibility study. Gastrointestinal endoscopy 2011;74(1):87–95 doi 10.1016/j.gie.2011.03.1235.

