## Supplemental Information for "A multi-analyte serum biomarker panel for early detection of pancreatic adenocarcinoma"

Supplementary Table S1: ELISA kits utilized.

| Analyte | Vendor | Part Number | Name |
|---|---|---|---|
| ALCAM | R&D Systems | DY656 | Human ALCAM DuoSet |
| ANG | R&D Systems | DY265 | Human Angiogenin DuoSet |
| AXL | R&D Systems | DY154 | Human Axl DuoSet |
| BSG | R&D Systems | DY972 | Human EMMPRIN/CD147 DuoSet |
| CA 19-9 | Diagnostic Automation | 6909-16 | Human CA-19-9 ELISA Test Kit |
| CEA | ALPCO | 25-CEAHU-E01 | Carcinoembryonic Antigen ELISA (CEA ELISA) |
| CEACAM1 | R&D Systems | DY2244 | Human CEACAM-1/CD66a DuoSet |
| COL18A1 | R&D Systems | DY1098 | Human Endostatin DuoSet |
| EPCAM | R&D Systems | DY960 | Human EpCAM/TROP1 Duoset |
| HA | R&D Systems | DY3614-05 | Hyaluronan DuoSet |
| HP | R&D Systems | DHAPG0 | Human Haptoglobin Quantikine ELISA Kit |
| HP | Aviva Systems Biology | OKIA00064 | Haptoglobin ELISA Kit (Human) |
| ICAM1 | R&D Systems | DY720 | Human ICAM-1/CD54 DuoSet |
| IGFBP2 | R&D Systems | DY674 | Human IGFBP-2 DuoSet |
| IGFBP4 | R&D Systems | DY804 | Human IGFBP-4 DuoSet |
| LCN2 | R&D Systems | DY1757 | Human Lipocalin-2/NGAL DuoSet |
| LRG1 | US Biological | USB 026489 | Leucine Rich Alpha-2-Glycoprotein 1 (LRG1) BioAssay ELISA Kit (Human) |
| LRG1 | Aviva Systems Biology | OKEH01377 | LRG1 ELISA Kit (Human) |
| MMP2 | R&D Systems | DY902 | Human MMP-2 DuoSet |
| MMP7 | R&D Systems | DY907 | Human Total MMP-7 DuoSet |
| MMP9 | R&D Systems | DY911 | Human MMP-9 DuoSet |
| MSLN | R&D Systems | DY3265 | Human Mesothelin Propeptide/MPF DuoSet |
| PARK7 | R&D Systems | DY3995 | Human Park7/DJ-1 DuoSet |
| PPBP | R&D Systems | DY393 | Human CXCL7/NAP-2 Duoset |
| PRG4 | US Biological | USB 027755 | Proteoglycan 4 (PRG4) BioAssay ELISA Kit (Human) |
| PRG4 | Aviva Systems Biology | OKEH00681 | PRG4 ELISA Kit (Human) |
| SPARCL1 | R&D Systems | DY2728 | Human SPARC-like 1/SPARCL1 DuoSet |
| SPP1 | R&D Systems | DY1433 | Human Osteopontin (OPN) DuoSet |
| TGFBI | R&D Systems | DY2935 | Human beta IG-H3 DuoSet |
| THBS1 | R&D Systems | DY3074 | Human Thrombospondin-1 DuoSet |
| TIMP1 | R&D Systems | DY970 | Human TIMP-1 DuoSet |
| TNFRSF1A | R&D Systems | DY225 | Human sTNF RI/TNFRSF1A DuoSet |
| VEGFC | R&D Systems | DY752B | Human VEGF-C DuoSet |

Supplementary Table S2: Analytes previously identified as potential biomarkers in early stage PDAC cases, pre-diagnostic samples, and/or cases with PanIN lesions.

| Analyte | Name | Reference |
|---|---|---|
| ALCAM | activated leukocyte cell adhesion molecule | (1-3) |
| ANG | angiogenin | (3-6) |
| AXL | AXL receptor tyrosine kinase | (3, 7) |
| BAG3 | BCL2-associated athanogene 3 | (3, 8, 9) |
| BSG | basigin (EMMPRIN, CD147) | (3, 10-13) |
| CA 19-9 | cancer antigen 19-9 | (14, 15) |
| CCL24 | C-C motif chemokine ligand 24 (eotaxin-2) | (3, 16) |
| CEA | carcinoembryonic antigen | (15, 17) |
| CEACAM1 | carcinoembryonic antigen-related cell adhesion molecule 1 | (3, 18, 19) |
| CLU | clusterin | (3, 20) |
| COL18A1 | collagen, type XVIII, alpha 1 (endostatin) | (2, 21) |
| EPCAM | epithelial cell adhesion molecule | (22) |
| HA | soluble hyaluronic acid | (23, 24) |
| HP | haptoglobin | (25, 26) |
| ICAM1 | intercellular adhesion molecule 1 | (2-4, 27) |
| IGFBP2 | insulin-like growth factor binding protein 2 | (3, 4, 28) |
| IGFBP4 | insulin-like growth factor binding protein 4 | (2, 3, 29) |
| LCN2 | lipocalin 2 (NGAL) | (2-4, 7) |
| LRG1 | leucine-rich alpha-2-glycoprotein 1 | (3, 4, 30) |
| MMP2 | matrix metallopeptidase 2 | (3, 4, 10, 31-33) |
| MMP7 | matrix metallopeptidase 7 | (3, 34-38) |
| MMP9 | matrix metallopeptidase 9 | (3, 33, 39, 40) |
| MSLN | mesothelin | (3, 28, 41-45) |
| PARK7 | DJ-1 protein | (3, 46, 47) |
| PF4 | platelet factor 4 | (4) |
| PPBP | platelet basic protein | (4) |
| PRG4 | proteoglycan 4 | (2, 4) |
| SPARCL1 | SPARC-like 1 (hevin) | (2, 4, 48) |
| SPP1 | secreted phosphoprotein 1 (osteopontin) | (3, 34, 49-52) |
| TGFBI | transforming growth factor, beta-induced, 68kDa | (2, 4, 23) |
| THBS1 | thrombospondin 1 | (1, 2, 4, 40, 53) |
| TIMP1 | TIMP metallopeptidase inhibitor 1 | (2-4, 52, 54, 55) |
| TNFRSF1A | tumor necrosis factor receptor superfamily, member 1A | (2, 56) |
| VEGFC | vascular endothelial growth factor C | (3, 57) |

Supplementary Table S3: Pilot analyses for analytes not previously specifically evaluated in early stage PDAC (30-60 samples per group).

| | | CON vs. PDAC | | ChPT vs. PDAC | |
|---|---|---|---|---|---|
| | Name | AUC (95% CI) | *P* value* | AUC (95% CI) | *P* value* |
| Included Analytes | | | | | |
| CEACAM1 | carcinoembryonic antigen-related cell adhesion molecule 1 | 0.87 (0.80 - 0.93) | <0.0001 | 0.77 (0.68 - 0.85) | <0.0001 |
| HA | soluble hyaluronic acid | 0.78 (0.69 - 0.85) | <0.0001 | 0.66 (0.56 - 0.75) | 0.002 |
| AXL | AXL receptor tyrosine kinase | 0.76 (0.68 - 0.84) | <0.0001 | 0.67 (0.57 - 0.76) | 0.002 |
| IGFBP2 | insulin-like growth factor binding protein 2 | 0.76 (0.67 - 0.84) | <0.0001 | 0.55 (0.44 - 0.65) | 0.352 |
| HP | haptoglobin | 0.75 (0.63 - 0.86) | 0.001 | 0.57 (0.42 - 0.71) | 0.404 |
| LRG1 | leucine-rich alpha-2-glycoprotein 1 | 0.72 (0.63 - 0.81) | <0.0001 | 0.53 (0.42 - 0.65) | 0.562 |
| BAG3 | BCL2-associated athanogene 3 | 0.71 (0.63 - 0.80) | <0.0001 | 0.49 (0.37 - 0.61) | 0.816 |
| MMP9 | matrix metallopeptidase 9 | 0.70 (0.61 - 0.79) | <0.0001 | 0.57 (0.46 - 0.67) | 0.234 |
| BSG | basigin (EMMPRIN, CD147) | 0.64 (0.53 - 0.74) | 0.006 | 0.58 (0.47 - 0.69) | 0.133 |
| MMP2 | matrix metallopeptidase 2 | 0.64 (0.54 - 0.74) | 0.008 | 0.49 (0.39 - 0.60) | 0.918 |
| Excluded Analytes | | | | | |
| PF4 | platelet factor 4 | 0.60 (0.48 - 0.72) | 0.129 | 0.56 (0.42 - 0.70) | 0.416 |
| CCL24 | C-C motif chemokine ligand 24 (eotaxin-2) | 0.59 (0.42 - 0.76) | 0.356 | 0.51 (0.30 - 0.72) | 0.942 |
| CLU | clusterin | 0.50 (0.27 - 0.76) | 0.987 | nd | nd |

*by Wilcoxon rank sum test

Supplementary Table S4: Ontology analysis of selected analytes.

| Biological Process | Count | Fold Enrichment | FDR | Genes |
|---|---|---|---|---|
| embryo implantation | 4 | 59.2 | 0.00251 | MMP2, BSG, SPP1, MMP9 |
| extracellular matrix disassembly | 6 | 49.1 | 0.00004 | MMP7, MMP2, BSG, SPP1, TIMP1, MMP9 |
| collagen catabolic process | 4 | 38.9 | 0.00669 | COL18A1, MMP7, MMP2, MMP9 |
| platelet degranulation | 4 | 24.2 | 0.02425 | VEGFC, PPBP, TIMP1, THBS1 |
| cellular protein metabolic process | 4 | 21.1 | 0.03244 | IGFBP4, MMP2, IGFBP2, TGFBI |
| leukocyte migration | 4 | 20.4 | 0.03248 | CEACAM1, BSG, MMP9, ICAM1 |
| extracellular matrix organization | 6 | 19.0 | 0.00153 | COL18A1, BSG, SPP1, TGFBI, THBS1, ICAM1 |
| angiogenesis | 6 | 16.7 | 0.00172 | COL18A1, CEACAM1, MMP2, VEGFC, ANG, TGFBI |
| response to drug | 6 | 12.3 | 0.00537 | COL18A1, IGFBP2, LCN2, VEGFC, THBS1, ICAM1 |
| inflammatory response | 7 | 11.5 | 0.00172 | AXL, IGFBP4, SPP1, PPBP, PARK7, THBS1, TNFRSF1A |
| cell adhesion | 8 | 10.8 | 0.00095 | COL18A1, CEACAM1, ALCAM, SPP1, TGFBI, MSLN, THBS1, ICAM1 |
| Cellular Component | | | | |
| platelet alpha granule lumen | 4 | 49.1 | 0.00071 | VEGFC, PPBP, TIMP1, THBS1 |
| basement membrane | 3 | 25.6 | 0.04134 | COL18A1, TIMP1, TGFBI |
| proteinaceous extracellular matrix | 7 | 17.6 | 0.00003 | COL18A1, MMP7, MMP2, SPARCL1, TIMP1, TGFBI, MMP9 |
| membrane raft | 4 | 13.1 | 0.02521 | BSG, PARK7, ICAM1, TNFRSF1A |
| extracellular matrix | 5 | 11.4 | 0.00725 | COL18A1, MMP7, MMP2, TGFBI, THBS1 |
| extracellular space | 21 | 10.5 | 0.00000 | COL18A1, MMP7, IGFBP4, MMP2, IGFBP2, HP, VEGFC, PPBP, MSLN, THBS1, MMP9, ICAM1, TNFRSF1A, LRG1, AXL, SPP1, LCN2, SPARCL1, ANG, TIMP1, TGFBI |
| cell surface | 8 | 10.0 | 0.00011 | CEACAM1, MMP7, EPCAM, AXL, MSLN, THBS1, ICAM1, TNFRSF1A |
| extracellular region | 18 | 7.5 | 0.00000 | COL18A1, MMP7, IGFBP4, MMP2, IGFBP2, HP, VEGFC, PRG4, PPBP, THBS1, MMP9, TNFRSF1A, LRG1, SPP1, LCN2, ANG, TIMP1, TGFBI |
| extracellular exosome | 20 | 4.8 | 0.00000 | COL18A1, MMP7, IGFBP2, HP, PARK7, THBS1, MMP9, ICAM1, CEACAM1, LRG1, ALCAM, EPCAM, AXL, BSG, SPP1, LCN2, SPARCL1, ANG, TIMP1, TGFBI |
| Molecular Function | | | | |
| extracellular matrix binding | 4 | 96.2 | 0.00078 | SPP1, SPARCL1, TGFBI, THBS1 |
| Keyword | | | | |
| Collagen degradation | 3 | 127.0 | 0.00299 | MMP7, MMP2, MMP9 |
| Pyrrolidone carboxylic acid | 4 | 32.8 | 0.00299 | CEACAM1, EPCAM, LCN2, ANG |
| Extracellular matrix | 6 | 17.7 | 0.00033 | COL18A1, MMP7, MMP2, SPARCL1, TGFBI, MMP9 |
| Zymogen | 4 | 14.2 | 0.02765 | MMP7, MMP2, PARK7, MMP9 |
| Cell adhesion | 7 | 11.1 | 0.00041 | COL18A1, ALCAM, SPP1, TGFBI, MSLN, THBS1, ICAM1 |
| Secreted | 21 | 8.1 | 0.00000 | COL18A1, MMP7, IGFBP4, MMP2, IGFBP2, HP, VEGFC, PRG4, PPBP, MSLN, MMP9, TNFRSF1A, CEACAM1, LRG1, ALCAM, SPP1, LCN2, SPARCL1, ANG, TIMP1, TGFBI |

| | | | | |
|---|---|---|---|---|
| Immunoglobulin domain | 5 | 7.0 | 0.04313 | CEACAM1, ALCAM, AXL, BSG, ICAM1 |
| Disulfide bond | 24 | 5.3 | 0.00000 | COL18A1, IGFBP4, MMP2, IGFBP2, HP, VEGFC, PRG4, PPBP, MSLN, THBS1, MMP9, ICAM1, TNFRSF1A, CEACAM1, LRG1, ALCAM, EPCAM, AXL, BSG, LCN2, SPARCL1, ANG, TIMP1, TGFBI |
| Signal | 26 | 4.8 | 0.00000 | COL18A1, HP, MSLN, THBS1, ICAM1, ALCAM, EPCAM, BSG, SPP1, TIMP1, MMP7, IGFBP4, MMP2, IGFBP2, VEGFC, PRG4, PPBP, MMP9, TNFRSF1A, CEACAM1, LRG1, AXL, LCN2, SPARCL1, ANG, TGFBI |
| Glycoprotein | 22 | 3.7 | 0.00000 | COL18A1, IGFBP4, MMP2, IGFBP2, HP, VEGFC, PRG4, MSLN, THBS1, MMP9, ICAM1, TNFRSF1A, CEACAM1, LRG1, ALCAM, EPCAM, AXL, BSG, SPP1, LCN2, SPARCL1, TIMP1 |
| Polymorphism | 23 | 1.5 | 0.04313 | COL18A1, MMP7, IGFBP4, MMP2, IGFBP2, HP, PRG4, PARK7, MSLN, THBS1, MMP9, ICAM1, TNFRSF1A, CEACAM1, LRG1, ALCAM, EPCAM, AXL, BSG, SPP1, SPARCL1, ANG, TGFBI |

Supplementary Table S5: Diagnostic performance of individual analytes in the development data set.

| | CON vs. PDAC | | IPMN vs. PDAC | | ChPT vs. PDAC | |
|---|---|---|---|---|---|---|
| Analyte | AUC (95% CI) | *P* value | AUC (95% CI) | *P* value | AUC (95% CI) | *P* value |
| ALCAM | 0.71 (0.66 - 0.75) | **<0.0001** | 0.65 (0.58 - 0.71) | **<0.0001** | 0.59 (0.52 - 0.66) | **0.016** |
| ANG | 0.50 (0.45 - 0.55) | 0.921 | 0.52 (0.45 - 0.59) | 0.571 | 0.52 (0.45 - 0.59) | 0.664 |
| AXL | 0.68 (0.63 - 0.73) | **<0.0001** | 0.63 (0.56 - 0.69) | **0.001** | 0.56 (0.49 - 0.63) | 0.100 |
| BAG3 | 0.61 (0.56 - 0.66) | **<0.0001** | 0.52 (0.44 - 0.58) | 0.663 | 0.51 (0.44 - 0.59) | 0.742 |
| BSG | 0.59 (0.54 - 0.64) | **0.001** | 0.55 (0.48 - 0.62) | 0.180 | 0.60 (0.52 - 0.67) | **0.007** |
| CA19-9 | 0.84 (0.80 - 0.89) | **<0.0001** | 0.79 (0.74 - 0.84) | **<0.0001** | 0.78 (0.72 - 0.83) | **<0.0001** |
| CEA | 0.69 (0.64 - 0.74) | **<0.0001** | 0.60 (0.53 - 0.67) | **0.006** | 0.57 (0.50 - 0.64) | **0.047** |
| CEACAM1 | 0.81 (0.77 - 0.85) | **<0.0001** | 0.82 (0.77 - 0.87) | **<0.0001** | 0.67 (0.61 - 0.74) | **<0.0001** |
| COL18A1 | 0.57 (0.51 - 0.61) | **0.010** | 0.60 (0.53 - 0.67) | **0.006** | 0.52 (0.44 - 0.59) | 0.637 |
| EPCAM | 0.55 (0.50 - 0.60) | **0.033** | 0.49 (0.41 - 0.56) | 0.693 | 0.59 (0.52 - 0.66) | **0.013** |
| HA | 0.64 (0.59 - 0.69) | **<0.0001** | 0.60 (0.54 - 0.66) | **0.006** | 0.51 (0.44 - 0.58) | 0.774 |
| HP | 0.63 (0.58 - 0.68) | **<0.0001** | 0.63 (0.55 - 0.69) | **0.001** | 0.59 (0.52 - 0.66) | **0.010** |
| ICAM1 | 0.80 (0.76 - 0.85) | **<0.0001** | 0.77 (0.71 - 0.82) | **<0.0001** | 0.58 (0.51 - 0.64) | **0.035** |
| IGFBP2 | 0.66 (0.61 - 0.71) | **<0.0001** | 0.65 (0.59 - 0.72) | **<0.0001** | 0.58 (0.51 - 0.65) | **0.025** |
| IGFBP4 | 0.61 (0.56 - 0.65) | **<0.0001** | 0.50 (0.43 - 0.58) | 0.922 | 0.49 (0.42 - 0.56) | 0.830 |
| LCN2 | 0.57 (0.52 - 0.61) | **0.009** | 0.51 (0.44 - 0.59) | 0.724 | 0.56 (0.47 - 0.63) | 0.126 |
| LRG1 | 0.59 (0.54 - 0.64) | **<0.0001** | 0.46 (0.40 - 0.53) | 0.336 | 0.53 (0.46 - 0.60) | 0.483 |
| MMP2 | 0.53 (0.48 - 0.58) | 0.176 | 0.52 (0.45 - 0.60) | 0.553 | 0.52 (0.44 - 0.59) | 0.657 |
| MMP7 | 0.70 (0.66 - 0.75) | **<0.0001** | 0.68 (0.61 - 0.74) | **<0.0001** | 0.62 (0.56 - 0.69) | **0.001** |
| MMP9 | 0.56 (0.51 - 0.60) | **0.028** | 0.53 (0.46 - 0.60) | 0.387 | 0.53 (0.45 - 0.61) | 0.394 |
| MSLN | 0.66 (0.62 - 0.71) | **<0.0001** | 0.57 (0.50 - 0.64) | 0.056 | 0.51 (0.43 - 0.58) | 0.768 |
| PARK7 | 0.58 (0.54 - 0.63) | 0.219 | 0.57 (0.50 - 0.63) | **0.010** | 0.60 (0.52 - 0.67) | **0.006** |
| PPBP | 0.58 (0.53 - 0.63) | **0.001** | 0.55 (0.47 - 0.62) | 0.064 | 0.58 (0.51 - 0.65) | **0.008** |
| PRG4 | 0.59 (0.54 - 0.63) | **0.002** | 0.57 (0.50 - 0.63) | 0.210 | 0.54 (0.46 - 0.61) | **0.028** |
| SPARCL1 | 0.77 (0.73 - 0.82) | **0.001** | 0.76 (0.70 - 0.82) | 0.071 | 0.54 (0.47 - 0.61) | 0.298 |
| SPP1 | 0.53 (0.48 - 0.58) | **<0.0001** | 0.59 (0.52 - 0.67) | **<0.0001** | 0.60 (0.53 - 0.67) | 0.251 |
| TGFBI | 0.61 (0.55 - 0.66) | **<0.0001** | 0.71 (0.65 - 0.77) | **<0.0001** | 0.61 (0.55 - 0.68) | **0.001** |
| THBS1 | 0.52 (0.47 - 0.56) | 0.533 | 0.56 (0.48 - 0.62) | 0.130 | 0.70 (0.63 - 0.76) | **<0.0001** |
| TIMP1 | 0.64 (0.59 - 0.69) | **<0.0001** | 0.65 (0.58 - 0.72) | **<0.0001** | 0.56 (0.49 - 0.62) | 0.117 |
| TNFRSF1A | 0.76 (0.71 - 0.80) | **<0.0001** | 0.63 (0.56 - 0.70) | **<0.0001** | 0.57 (0.50 - 0.64) | 0.061 |
| VEGFC | 0.54 (0.49 - 0.58) | 0.141 | 0.63 (0.56 - 0.69) | **0.001** | 0.47 (0.40 - 0.54) | 0.357 |

Supplementary Table S5: (continued)

| | CON vs. IPMN | | CON vs. ChPT | | IPMN vs. ChPT | |
|---|---|---|---|---|---|---|
| Analyte | AUC (95% CI) | *P* value | AUC (95% CI) | *P* value | AUC (95% CI) | *P* value |
| ALCAM | 0.59 (0.53 - 0.65) | **0.005** | 0.61 (0.55 - 0.68) | **<0.0001** | 0.55 (0.47 - 0.64) | 0.194 |
| ANG | 0.48 (0.42 - 0.55) | 0.595 | 0.52 (0.45 - 0.58) | 0.587 | 0.53 (0.46 - 0.61) | 0.431 |
| AXL | 0.58 (0.52 - 0.63) | **0.017** | 0.62 (0.56 - 0.68) | **<0.0001** | 0.55 (0.47 - 0.63) | 0.252 |
| BAG3 | 0.60 (0.54 - 0.66) | **0.002** | 0.59 (0.53 - 0.66) | **0.003** | 0.50 (0.42 - 0.59) | 0.964 |
| BSG | 0.54 (0.48 - 0.60) | 0.233 | 0.51 (0.44 - 0.57) | 0.787 | 0.55 (0.46 - 0.62) | 0.275 |
| CA19-9 | 0.63 (0.57 - 0.69) | **<0.0001** | 0.61 (0.54 - 0.68) | **0.001** | 0.50 (0.42 - 0.58) | 0.960 |
| CEA | 0.62 (0.56 - 0.68) | **<0.0001** | 0.62 (0.55 - 0.68) | **<0.0001** | 0.52 (0.44 - 0.60) | 0.670 |
| CEACAM1 | 0.52 (0.46 - 0.59) | 0.510 | 0.65 (0.59 - 0.71) | **<0.0001** | 0.67 (0.60 - 0.75) | **<0.0001** |
| COL18A1 | 0.53 (0.47 - 0.60) | 0.292 | 0.54 (0.48 - 0.61) | 0.163 | 0.57 (0.49 - 0.66) | 0.079 |
| EPCAM | 0.56 (0.50 - 0.63) | 0.058 | 0.64 (0.58 - 0.71) | **<0.0001** | 0.57 (0.49 - 0.65) | 0.099 |
| HA | 0.56 (0.50 - 0.61) | 0.077 | 0.66 (0.59 - 0.72) | **<0.0001** | 0.62 (0.54 - 0.70) | **0.005** |
| HP | 0.50 (0.44 - 0.57) | 0.938 | 0.73 (0.67 - 0.79) | **<0.0001** | 0.73 (0.65 - 0.80) | **<0.0001** |
| ICAM1 | 0.57 (0.51 - 0.63) | **0.023** | 0.75 (0.69 - 0.81) | **<0.0001** | 0.70 (0.63 - 0.78) | **<0.0001** |
| IGFBP2 | 0.53 (0.48 - 0.59) | 0.292 | 0.71 (0.65 - 0.77) | **<0.0001** | 0.70 (0.62 - 0.78) | **<0.0001** |
| IGFBP4 | 0.61 (0.54 - 0.67) | **0.001** | 0.62 (0.55 - 0.68) | **<0.0001** | 0.51 (0.42 - 0.59) | 0.905 |
| LCN2 | 0.59 (0.53 - 0.64) | **0.007** | 0.61 (0.54 - 0.68) | **0.001** | 0.55 (0.46 - 0.63) | 0.244 |
| LRG1 | 0.57 (0.50 - 0.63) | **0.046** | 0.56 (0.50 - 0.63) | **0.045** | 0.50 (0.42 - 0.59) | 0.944 |
| MMP2 | 0.55 (0.49 - 0.62) | 0.106 | 0.54 (0.47 - 0.61) | 0.250 | 0.49 (0.40 - 0.57) | 0.845 |
| MMP7 | 0.52 (0.45 - 0.59) | 0.567 | 0.58 (0.52 - 0.64) | **0.012** | 0.56 (0.47 - 0.63) | 0.185 |
| MMP9 | 0.60 (0.54 - 0.66) | **0.002** | 0.49 (0.42 - 0.56) | 0.650 | 0.56 (0.48 - 0.64) | 0.155 |
| MSLN | 0.62 (0.57 - 0.68) | **<0.0001** | 0.64 (0.58 - 0.70) | **<0.0001** | 0.54 (0.46 - 0.63) | 0.289 |
| PARK7 | 0.54 (0.49 - 0.59) | 0.055 | 0.48 (0.41 - 0.54) | **0.032** | 0.56 (0.48 - 0.64) | 0.926 |
| PPBP | 0.53 (0.46 - 0.60) | 0.209 | 0.67 (0.61 - 0.73) | 0.493 | 0.63 (0.54 - 0.70) | 0.154 |
| PRG4 | 0.55 (0.50 - 0.60) | 0.394 | 0.54 (0.48 - 0.61) | **<0.0001** | 0.51 (0.43 - 0.59) | **0.002** |
| SPARCL1 | 0.48 (0.42 - 0.55) | 0.152 | 0.81 (0.76 - 0.86) | 0.161 | 0.79 (0.73 - 0.86) | 0.740 |
| SPP1 | 0.56 (0.50 - 0.62) | 0.577 | 0.57 (0.50 - 0.63) | **<0.0001** | 0.50 (0.42 - 0.58) | **<0.0001** |
| TGFBI | 0.63 (0.57 - 0.68) | **<0.0001** | 0.51 (0.45 - 0.58) | 0.663 | 0.59 (0.51 - 0.68) | **0.025** |
| THBS1 | 0.52 (0.46 - 0.58) | 0.593 | 0.70 (0.64 - 0.76) | **<0.0001** | 0.75 (0.68 - 0.82) | **<0.0001** |
| TIMP1 | 0.53 (0.46 - 0.60) | 0.388 | 0.58 (0.51 - 0.65) | **0.011** | 0.60 (0.51 - 0.67) | **0.020** |
| TNFRSF1A | 0.65 (0.59 - 0.71) | **<0.0001** | 0.64 (0.57 - 0.71) | **<0.0001** | 0.54 (0.45 - 0.62) | 0.356 |
| VEGFC | 0.58 (0.52 - 0.63) | **0.016** | 0.50 (0.44 - 0.56) | 0.983 | 0.58 (0.50 - 0.66) | 0.058 |

AUC = area under the ROC curve, CI = bootstrap confidence interval, *P* value by Wilcoxon rank sum test, bold text indicates $P < 0.05$

Supplementary Table S6: Variable Importance. Individual methods (top row) were applied to an intermediate subset of cases (74 CON, 60 ChPT, 76 IPMN, 122 PDAC) and the relative contribution of the individual predictors (analytes, age, gender) was calculated for the resulting discriminant algorithm. The highest contributing variable was assigned an importance of 100.

| Predictor | GLMnet | KNN | NNET | RF | SVM |
|---|---|---|---|---|---|
| Age | 39 | 35 | 57 | 9 | 35 |
| ALCAM | 10 | 67 | 41 | 15 | 67 |
| ANG | 0 | 1 | 38 | 5 | 1 |
| AXL | 0 | 56 | 30 | 6 | 56 |
| BAG3 | 0 | 8 | 17 | 5 | 8 |
| BSG | 0 | 23 | 36 | 4 | 23 |
| CA 19-9 | 78 | 100 | 100 | 100 | 100 |
| CEA | 9 | 31 | 40 | 8 | 31 |
| CEACAM1 | 100 | 96 | 32 | 64 | 96 |
| COL18A1 | 0 | 24 | 25 | 3 | 24 |
| EPCAM | 9 | 9 | 14 | 5 | 9 |
| Gender | 14 | 12 | 34 | 0 | 12 |
| HA | 0 | 53 | 64 | 9 | 53 |
| HP | 1 | 14 | 24 | 4 | 14 |
| ICAM1 | 28 | 70 | 15 | 19 | 70 |
| IGFBP2 | 0 | 39 | 44 | 5 | 39 |
| IGFBP4 | 16 | 20 | 32 | 3 | 20 |
| LCN2 | 6 | 15 | 19 | 5 | 15 |
| LRG1 | 0 | 0 | 33 | 5 | 0 |
| MMP2 | 11 | 13 | 69 | 7 | 13 |
| MMP7 | 7 | 54 | 22 | 7 | 54 |
| MMP9 | 0 | 8 | 23 | 5 | 8 |
| MSLN | 3 | 9 | 19 | 5 | 9 |
| PARK7 | 0 | 17 | 18 | 4 | 17 |
| PPBP | 0 | 6 | 42 | 4 | 6 |
| PRG4 | 8 | 11 | 62 | 5 | 11 |
| SPARCL1 | 43 | 29 | 43 | 11 | 29 |
| SPP1 | 11 | 70 | 41 | 14 | 70 |
| TGFBI | 17 | 42 | 45 | 7 | 42 |
| THBS | 0 | 1 | 39 | 6 | 1 |
| TIMP1 | 0 | 45 | 42 | 5 | 45 |
| TNFRSF1A | 0 | 47 | 0 | 4 | 47 |
| VEGFC | 0 | 1 | 35 | 3 | 1 |

GLMnet = lasso regression; KNN = k-nearest neighbors; NNET = neural network; RF = random forest; SVM = support vector machines, radial

Supplementary Table S7: Method Elimination. The effect of sequential elimination of methods from the ensemble model on diagnostic accuracy was evaluated using an intermediate subset of cases (74 CON, 60 ChPT, 76 IPMN, 122 PDAC). Cross-validated accuracy was determined maximizing area under the ROC curve. Kappa indicates Cohen's Kappa, a measure of classification accuracy that takes into account the possibility that agreement occurs by chance.

| SVM | KNN | GLMnet | RF | NNET | Kappa | Accuracy (95% CI) |
|---|---|---|---|---|---|---|
| + | + | + | + | + | 1 | 1 (0.989, 1) |
| | + | + | + | + | 1 | 1 (0.989, 1) |
| + | | + | + | + | 0.994 | 0.997 (0.983, 1) |
| + | + | | + | + | 1 | 1 (0.989, 1) |
| + | + | + | | + | 0.815 | 0.916 (0.880, 0.943) |
| + | + | + | + | | 1 | 1 (0.989, 1) |
| + | + | + | | | 0.785 | 0.904 (0.867, 0.933) |
| + | + | | + | | 1 | 1 (0.989, 1) |
| + | + | | | + | 1 | 1 (0.989, 1) |
| + | | + | + | | 1 | 1 (0.989, 1) |
| + | | + | | + | 0.670 | 0.849 (0.806, 0.886) |
| + | | | + | + | 1 | 1 (0.989, 1) |
| | + | + | + | | 1 | 1 (0.989, 1) |
| | + | + | | + | 0.563 | 0.810 (0.764, 0.851) |
| | + | | + | + | 1 | 1 (0.989, 1) |
| | | + | + | + | 1 | 1 (0.989, 1) |
| + | + | | | | 0.774 | 0.898 (0.860, 0.928) |
| + | | + | | | 0.682 | 0.855 (0.813, 0.891) |
| + | | | + | | 1 | 1 (0.989, 1) |
| + | | | | + | 0.994 | 0.997 (0.983, 1) |
| | + | + | | | 0.583 | 0.819 (0.774, 0.859) |
| | + | | + | | 1 | 1 (0.989, 1) |
| | + | | | + | 0.737 | 0.886 (0.846, 0.918) |
| | | + | + | | 1 | 1 (0.989, 1) |
| | | + | | + | 0.587 | 0.819 (0.774, 0.859) |
| | | | + | + | 1 | 1 (0.989, 1) |
